# Plasma protein prioritisation in rheumatoid arthritis reveals druggable targets and shared biology with cardiovascular diseases

**DOI:** 10.64898/2026.06.10.26355332

**Authors:** Saleh Alduhayhi, Andrew P Morris, Sizheng Steven Zhao, John Bowes

**Affiliations:** Centre for Genetics and Genomics Versus Arthritis, Centre for Musculoskeletal Research, Manchester Academic Health Science Centre, The University of Manchester, Manchester, United Kingdom; Public Health Department, College of Public Health, Imam Abdulrahman Bin Faisal University, Dammam, Saudi Arabia; Centre for Epidemiology Versus Arthritis, Centre for Musculoskeletal Research, Manchester Academic Health Science Centre, University of Manchester, Manchester, United Kingdom; NIHR Manchester Biomedical Research Centre, Central Manchester NHS Foundation Trust, Manchester Academic Health Science Centre, Manchester, United Kingdom

## Abstract

**Background:** Rheumatoid arthritis (RA) is an autoimmune inflammatory disease with complex and incompletely understood molecular mechanisms. Understanding circulating proteins associated with RA may improve understanding of disease biology and clarify its pathological links with cardiometabolic comorbidities.

**Methods:** A proteome-wide two-sample Mendelian randomisation (MR) drug target analysis was conducted using plasma proteins measured in 54,219 participants from the UK Biobank Pharma Proteomics Project as exposures and RA and cardiometabolic diseases as the outcomes. Summary statistics for RA included 53,663 cases and 1,070,200 controls. Colocalisation analysis was performed to confirm shared single causal variants and prioritise RA proteins supported by both MR and colocalisation. The prioritised proteins were then evaluated in the Accelerating Medicines Partnership RA Phase II synovial single-cell dataset for cell-type expression patterns. Druggability was then assessed followed by analysis of genetic overlap between RA-associated proteins and cardiometabolic diseases.

**Results:** 37 plasma proteins had a causal effect on RA risk, supported by combined evidence from MR and conditional colocalisation. In synovial tissue, TPPP3, RARRES2, AKAP12, and GGT5 were predominantly expressed in stromal and endothelial cell clusters. Druggability assessment identified IFNGR2, IL6R, CD40, and FCGR2B as Tier 1 targets. However, several biologically relevant proteins, including RARRES2, AKAP12, TPPP3, and SNX2, had limited available druggability data. Genetic overlap analysis demonstrated shared protein signals between RA and cardiovascular diseases, including overlap of RARRES2 and TPPP3 with coronary artery disease (CAD) and FCGR2B with atrial fibrillation (AF). To approximate the therapeutic effect of target inhibition, the direction of effect estimates for proteins showing overlap between RA-CAD and RA-AF was reversed.

**Conclusion:** This study identified circulating proteins involved in RA pathogenesis and reveals shared mechanisms between RA and cardiovascular diseases. While some proteins showed clear translational potential targets, several prioritised proteins had limited available druggability information and could not be confidently classified. Addressing these gaps may help identify new targets relevant to RA management. Future work should also use phenome-wide MR studies to evaluate potential on-target adverse effects of protein inhibition across RA-CAD and RA-AF.

## Introduction

Rheumatoid arthritis (RA) is a chronic systemic autoimmune disease characterised by persistent inflammation of the synovial joints.^1, 2, 3^ Synovial joints are highly specialised structures that preserve movement through coordinated homeostatic mechanisms while the synovium and its hyaluronan-rich synovial fluid help maintain lubrication. When this mechanism is disrupted, the synovium becomes inflamed leading to synovitis with activation of synovial lining cells, interstitial macrophages, endothelial cells, lymphocytes, and fibroblasts which in RA contributes to bone damage.^4, 5, 6^ Although RA is primarily defined by joint pathology, it is a systemic inflammatory condition with effects that extend beyond the synovium.^7^ Furthermore, RA is also associated with other cardiometabolic diseases which might worsen outcomes in patients with RA.^8, 9^

Circulating proteins in the blood are an important source of information in RA research because it reflects the systemic immune and inflammatory activity that characterise the disease. Circulating proteins, cytokines, immune cells, and autoantibodies can provide accessible molecular markers and offer insights into disease mechanism. For example, blood-based serology including the presence or absence of rheumatoid factor and anti-citrullinated protein antibodies contributes to RA classification and distinguishes seropositive from seronegative disease. However, blood-derived measures also have clear limitations. Since RA pathology is centred in the synovium, circulating protein levels may not fully represent the biological processes taking place in the target tissue where inflammation and structural damage develop. Signals observed in blood may reflect broader systemic inflammation rather than direct involvement in synovial pathology. ^10, 11, 12, 13^

Recent advances in large-scale genomic and proteomic data including genome-wides associations studies (GWASs) and the UK Biobank Pharma Proteomic Projects (UKB-PPP) have created new opportunities to investigate the molecular determinants of complex diseases. GWASs have identified numerous genetic loci associated with RA susceptibility while UKB-PPP provides extensive data on circulating protein levels derived from blood samples that can be integrated with genetic information to explore potential therapeutic targets.14, 15, 16, 17

Cis-Mendelian randomisation (MR) is an extension framework of genome-wide MR in which genetic variants located within or near the gene encoding a protein are used as instruments. Cis-acting variants are more likely to affect the encoded protein directly, and this approach has been increasingly adopted for drug target discovery as well as assessment of potential adverse effects.^18, 19, 20, 21, 22^

Despite the growing availability of drug target research using proteomic data, several important gaps remain in translating these findings into biologically meaningful and clinically relevant targets. There is a need to prioritise RA-associated proteins to assess their expression in synovial tissue, evaluate their druggability, and investigate their overlap with cardiometabolic diseases. Addressing these gaps may improve the interpretation of blood-based proteomic findings, strengthen target prioritisation for RA, and provide broader insight into the molecular relationships between RA and related comorbid conditions.

## Method

### Study design

This study employed a multi-step analytical strategy to identify and characterise proteins involved in RA. First, Two-sample MR was performed using plasma proteins as exposures and RA as the outcome to identify proteins with potential causal effects on disease risk. Then, colocalisation analysis was applied to determine whether the protein and RA association signals were driven by the same underlying causal variants. Proteins supported by both MR and colocalisation were prioritised for further investigation. The biological relevance of these prioritised proteins was assessed in synovial tissue using the Accelerating Medicines Partnership (AMP) RA Phase II across major cell populations. Then, their druggable potential was evaluated using druggable tier classifications and Open Targets data. Finally, the involvement of these proteins in cardiometabolic disease risk was further examined to identify potential shared mechanisms between RA and cardiometabolic comorbidities (Figure 1).

**Figure 1.**
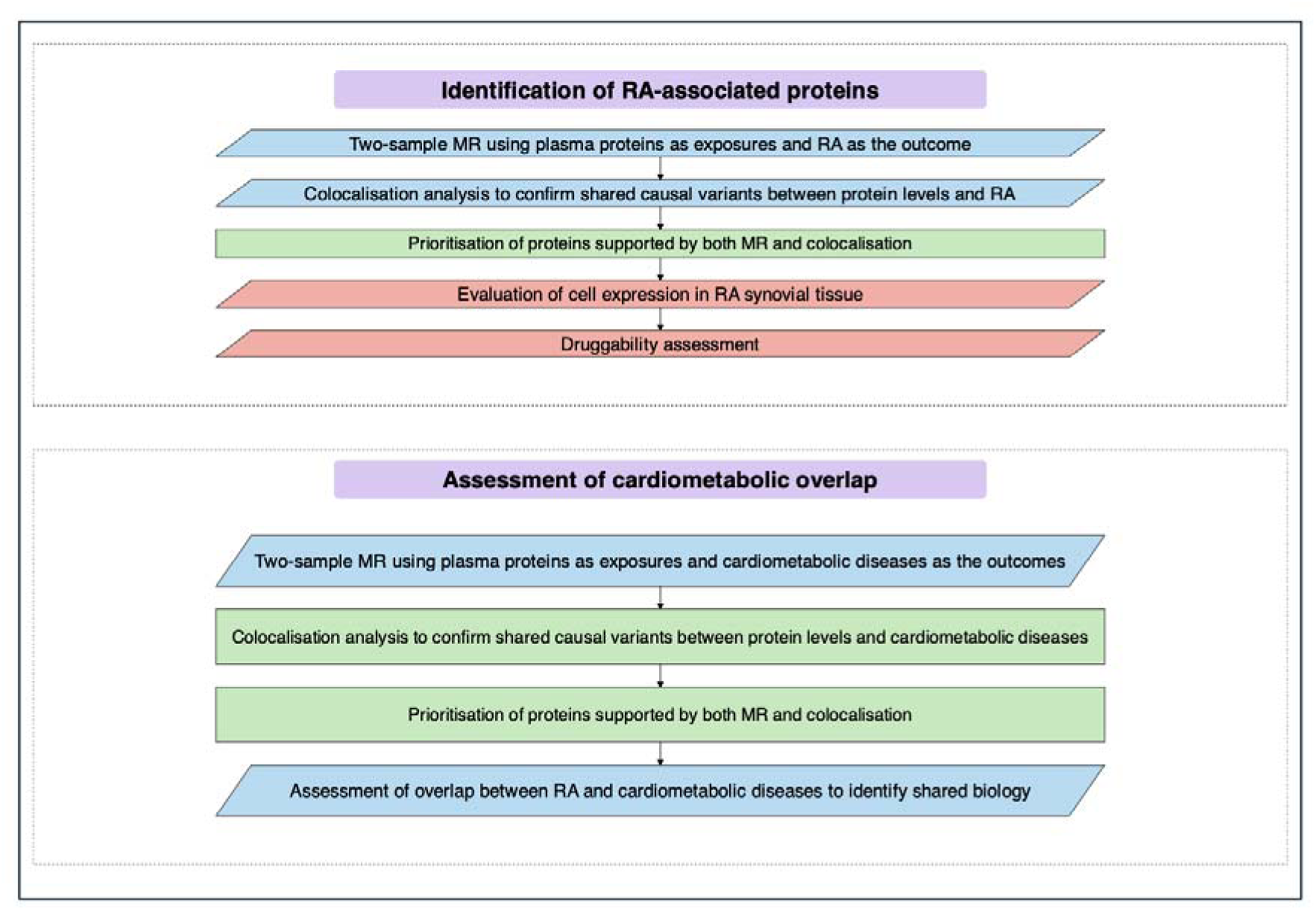
Demonstration of the analytical workflow used to identify and characterise proteins associated with RA and their shared biological effects with cardiometabolic diseases.

### Study population

The present study used proteomic data generated from the UKB-PPP. The dataset included 54,219 participants with available plasma proteomic measurements. Proteomic profiling was performed on blood plasma samples using antibody-based Olink Explore 3072 proximity extension assay, quantifying 2,941 analytes corresponding to 2,923 unique proteins across multiple biological domains, including cardiometabolic, inflammatory, neurological, and oncological pathways.^16^

Corrected meta-analysis due to sample overlap was performed by combining summary statistics from two large GWASs of RA using METACARPA,^23, 24, 25^ which accounts for unknown sample overlap. The combined dataset comprised 53,663 RA cases and 1,070,200 controls. The first GWAS, conducted by Saevarsdottir et al. included 31,313 cases and 995,377 controls from multiple European cohorts.^14^ RA cases were identified through rheumatologist diagnosis, national Scandinavian rheumatology quality registries and/or registry ICD-10 codes. The second GWAS was performed by Ishigaki et al. analysing 22,350 cases and 74,823 controls. RA cases met the 1987 classification criteria or the 2010 American College of Rheumatology/European League Against Rheumatism criteria or were diagnosed by a rheumatologist.^15^

GWAS summary statistics for cardiometabolic diseases were obtained from large previously published meta-analyses. Specifically, hypertension data was obtained from Zhu et al. (144,793 cases and 313,761 controls)^26^, coronary artery disease (CAD) from Aragam et al. (181,522 cases and 984,168 controls),^27^ atrial fibrillation (AF) from Nielsen et al. (60,620 cases and 970,216 controls),^28^ heart failure (HF) from Shah et al. (47,309 cases and 930,014 controls),^29^ stroke from Mishra et al. (73,652 cases and 1,234,808 controls),^30^ and type two diabetes mellitus (T2DM) from Mahajan et al. (80,154 cases and 853,816 controls).^31^ Detailed definitions of each cardiometabolic disease phenotype are provided in Supplementary Table 1.

### Combining evidence from MR drug-target and colocalisation

Cis-MR analyses were performed to evaluate the causal effects of genetically predicted perturbations of plasma protein levels from the UKB-PPP on RA and cardiometabolic disease risk. Genetic instruments were selected within ±500 KB window of the encoding gene to capture cis-acting variation. To balance statistical power and bias due to linkage disequilibrium (LD), LD clumping using threshold of *r*^2^ < 0.1 within a 1000 kb window was applied, keeping variants with *P* < 5×10-8.^32^ Exposure outcome summary statistics were harmonised to ensure consistent alignment of effect alleles. SNPs with reversed alleles were corrected by flipping the direction of the SNP-outcome effect. Palindromic SNPs were also excluded from the analysis.^33^ For exposures instrumented by a single SNP, causal effects were estimated using the Wald ratio method.^34^ For exposures with multiple independent SNPs, causal estimates were derived using the inverse variance weighted (IVW) method, which combines SNP-specific Wald ratios within a meta-analysis framework.^35^ False Discovery Rate (FDR) was applied to account for multiple testing and statistical significance was defined as *P*_FDR_<0.05.

To distinguish true shared causal signals from pleiotropy or confounding due to LD, the MR results were complemented by colocalisation. This approach assess whether the same genetic variant underlies associations in both exposure and outcome. Approximate Bayes factor framework was implemented using *coloc* R package to evaluate five hypotheses: no association with either trait (H_0_), association with only one trait (H_1_ or H_2_), association with both traits driven by distinct causal variants (H_3_), or association with both traits driven by a shared single causal variant (H_4_). Posterior probabilities were calculated using SNP-level effect sizes, standard errors, sample sizes, with prior probabilities set to P1=1×10-4, P2=1×10-4, and P12=1×10-5).^36^

In addition to the standard colocalisation, a conditional framework was applied to account for loci with weak outcome association of H_4_ due to limited statistical power rather than absence of a single shared causal variant. To address this, conditional probability of colicalisation was assessed using P(H4) / [P(H3) + P(H4)], representing the probability of a shared causal variant given the second trait has a causal variant in the region. Strong evidence of colocalisation was defined based on the conditional colocalisation (CC) as ≥ 70%.^37^

### Evaluation of cell expression in RA synovial tissue

To assess whether the prioritised plasma protein targets were also supported at the site of the disease, the expression of their corresponding encoding genes was examined in RA synovial tissue, using the AMP RA Phase II single-cell platform available through Immunogenomics.io. This platform is based on the synovial atlas reported by Zhang et al. and profiles of cells from the inflamed RA synovium across six major cell compartments: natural killer (NK) cells, T cells, myeloid cells, B cells, endothelial cells (E), and stromal cells (F). Expression was assessed primarily using the avg metric, which represents the mean Log2 Counts Per Million (log2CPM) of a gene within a given cell cluster. In this context, a higher avg value indicated stronger average expression of the corresponding gene in that synovial cell cluster.^38^ Prioritised plasma proteins were considered meaningfully expressed in synovial tissue when they showed statistically significant expression after multiple-testing correction (FDR < 0.05).

Protein expression estimates were then visualised using a heatmap, and a hierarchical clustering was applied to group columns in a way that allow visualisation of all protein expression patterns per each cell type cluster.

### Druggability assessment

Prioritised protein targets of RA were assessed further for druggability. First, targets were classified into tiers using the framework proposed by Finan et al. in the *Druggable Genome and Support for Target Identification and Validation in Drug Development*. In this framework, Tier 1 comprises genes encoding targets of approved drugs and clinical phase therapeutics, representing the strongest level of clinical validation. Tier 2 includes genes encoding proteins with known bioactive drug-like small molecule interactions or high sequence similarity to established drug targets, indicating likely tractability. Tier 3 includes genes encoding secreted or extracellular proteins, with more distant similarity to known drug targets, and members of key druggable protein families not included in Tier 1 and 2, reflecting potential but less well-established druggability.^39^

Additional therapeutic evidence was then obtained from the Open Targets Platform (an integrated database that combines genetic genomic and clinical evidence to help identify and prioritise potential drug targets, https://platform.opentargets.org). Data were mainly reviewed from the Target dataset for core gene/protein annotation, the Target Prioritisation dataset for target-specific prioritisation attributes, and the Clinical Indication dataset for approved and investigational drug indications. These datasets were used to extract drug pharmacological action, the maximum clinical stage reached by the target across all indications, and the maximum clinical stage for each drug-disease indication.^40^

## Result

### RA-prioritised proteins

A total of 343 circulating plasma proteins showed FDR-significant causal effects on RA risk, of which 37 circulating plasma proteins showed additional evidence by conditional colocalisation analysis. Of the 37 proteins, 19 proteins showed positive causal effects, indicating that genetically predicted increases in their circulating protein levels were associated with increased risk of RA while the remaining 18 proteins showed inverse causal effects. (Figure 2, Supplementary Table 2 shows all proteins < 0.05 FDR with their colocalisation and conditional colocalisation estimates).

**Figure 2.**
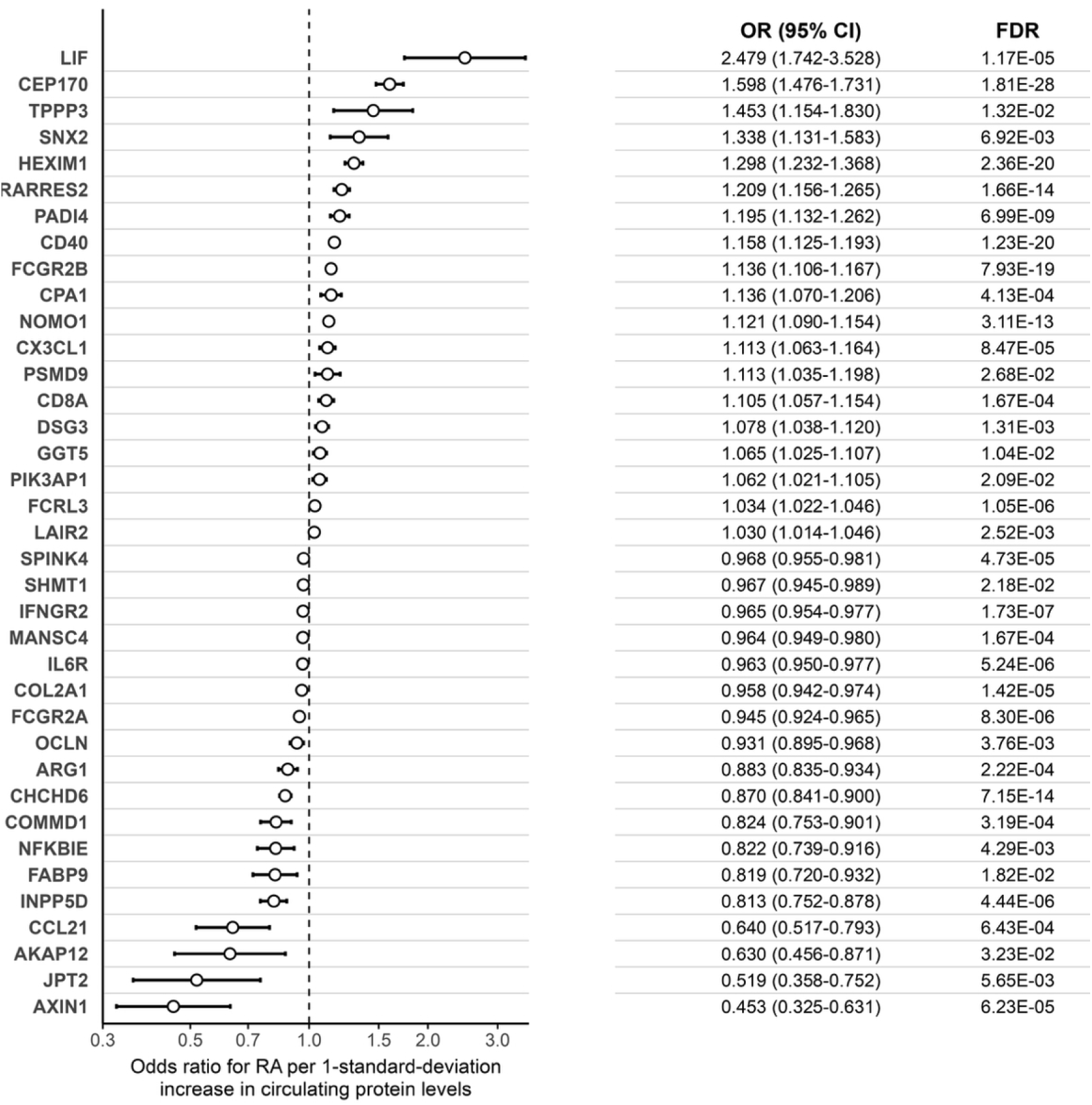
Forest plot showing the association between circulating protein levels (per 1-standard-deviation increase) and RA risk. Odds ratio (OR) were derived from IVW method and probability (*P)* values were corrected using FDR. Conditional colocalisation estimates of these proteins are found in Supplementary table 2

### Cell-type-specific expression of the prioritised proteins in RA synovial tissue

All the 37 RA-associated proteins showed statistically significant expression in at least one cell cluster except Fatty Acid-Binding Protein 9 (FABP9) which showed no significant expression across any cluster. Interleukin 6 Receptor (IL6R) was the protein that showed significant expression across most cell-type clusters (Supplementary Table 3).

The strongest recurrent pattern was observed for Fc Fragment of IgG, Low Affinity IIa (FCGR2A) and Fc fragment of IgG receptor IIb (FCGR2B), which were predominately\ expressed across multiple myeloid clusters. Cluster of Differentiation 40 (CD40) showed a broader immune distribution, being detected in both myeloid and B-cell clusters, with the highest expression in the B-4 cluster. T-cell surface glycoprotein CD8 alpha chain (CD8A) showed highly restricted pattern, being detected mainly across several T-cell clusters while leukocyte-associated immunoglobulin-like receptor 2 (LAIR2) was limited to T_Cells7 and NK-0 clusters. In contrast, Tubulin Polymerization-Promoting Protein Family Member 3 (TPPP3), Retinoic Acid Receptor Responder 2 (RARRES2), and A-Kinase Anchoring Protein 12 (AKAP12) were predominately observed in stromal and endothelial clusters. Gamma-glutamyltransferase 5 (GGT5) was significantly expressed in stromal cell clusters (F-5 and F-7). Chemokine (C-C motif) ligand 21(CCL21) showed a highly restricted signal limited to an endothelial cluster (E-4) but it was highly expressed, whereas sorting nexin 2 (SNX2) was detected across several clusters of myeloid, B and endothelial cells. Overall, these results indicated that the proteins identified from blood-based proteomic data have distinct synovial expression patterns, with most consistent signal centred on myeloid clusters (Figure 3, Supplementary Table 4 showed all estimates related to the 37 proteins).

**Figure 3.**
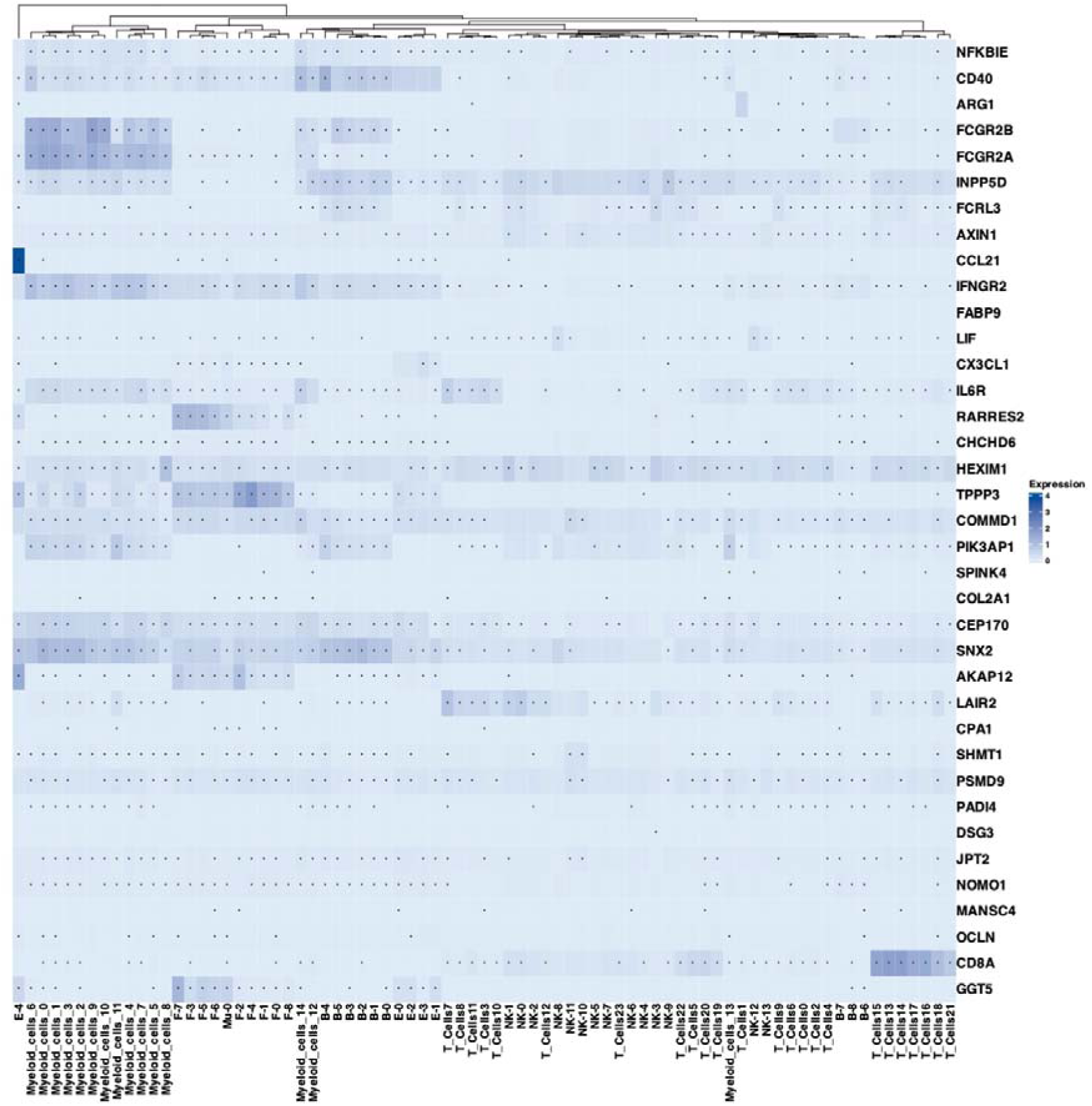
Heatmap of average expression levels of the RA prioritised proteins across synovial cell clusters in the AMP RA Phase II data. Rows represent proteins and columns represent synovial cell clusters. Colour intensity corresponds to the average expression level within each cluster. **F** = Stromal cells, **NK** = natural killer, **E** = endothelial cells. The asterisks (*) inside each cell denote statistically significant associations based on FDR-adjusted p-values < 0.05.

### Druggability assessment

Druggability assessment of the RA prioritised proteins showed that a subset already has evidence of therapeutic tractability. Interferon Gamma Receptor 2 (IFNGR2), IL6R, CD40, and FCGR2B were classified as Teir 1 targets, indicating the strongest level of existing drug-target support. However, the extent of clinical development differed across these targets.

IFNGR2 and IL6R each has a maxClinicalStage score (this score is based on the most advanced trial or approval stage reached for a target in any disease) of 1 indicating that these targets had reached the most advanced level of clinical development in at least one indication, whereas FCGR2B and CD40 had lower scores of 0.7 and 0.2 respectively indicating comparatively less advanced overall clinical progression. Notably, IL6R had approved use in RA (tocilizumab) while CD40 and FCGR2B showed more limited RA-specific development, reaching phase 2 (iscalimab) and phase ½ (obexelimab) respectively.

Several other proteins including RARRES2, AKAP12, TPPP3, and SNX2 had no much available druggable data, suggesting that they are currently not recognised as established drug targets (Supplementary Table 5 - 6). Together, these findings indicated that while some highly expressed proteins have clear translational potential, others remain less developed from a druggability perspective.

### Assessment of cardiometabolic overlap

Prioritised RA-associated proteins were examined in their potential overlap with the risk of the cardiometabolic diseases. The analysis identified overlapping proteins; RARRES2 and TPPP3 were overlapping across RA and CAD, and FCGR2B in RA and AF. In the original effect direction, genetically predicted higher levels of RARRES2 and TPPP3 were associated with increased risk of RA (RARRES2-RA, OR = 1.21 (95% confidence interval (CI) 1.16-1.27, *P*_FDR_ = 1.66×10-14, CC = 95%) and CAD (RARRES2-CAD, OR = 1.09 (95% CI 1.06-1.12, *P*_FDR_ = 1.46×10-10, CC = 76%), (TPPP3-RA, OR = 1.45 95% CI 1.15-1.83, *P*_FDR_ = 1.32 ×10-02, CC = 90%), and (TPPP3-CAD, OR = 1.31, 95% CI 1.13-1.52, *P*_FDR_ = 4.17×10-03, CC = 96%). Genetically predicted higher levels of FCGR2B were associated with increased risk of RA and AF. The corresponding estimates of FCGR2B-RA and FCGR2B-AF were (OR = 1.14, CI 1.11-1.17, *P*_FDR_ = 7.93×10-19, CC = 98%), and (OR = 1.04 CI 1.01 - 1.06, *P*_FDR_ = 1.45×10-02, CC = 93%), respectively.

To aid therapeutic interpretation, these estimates were expressed in the opposite biological direction. Thus, the ORs for RARRES2-RA, RARRES2-CAD, TPPP3-RA, and TPPP3-CAD were 0.83 (95% CI 0.79 - 0.87), 0.69 (95% CI 0.55 -0.87), 0.92 (95% CI 0.90- 0.94), and 0.76 (95% CI 0.66-0.89), respectively while FCGR2B-RA and FCGR2B-AF were 0.88 (95% CI 0.86-0.90), 0.96 (95% CI 0.94-0.99), respectively. These findings suggested that inhibition of these proteins might be associated with decreased risk in RA, CAD and AF (Figure 4 - 5, Supplementary Table 7 - 12 list the MR, colocalisation and conditional colocalisation estimates of the genetic predisposition to the plasma proteins and the risk of the cardiometabolic diseases).

**Figure 4.**
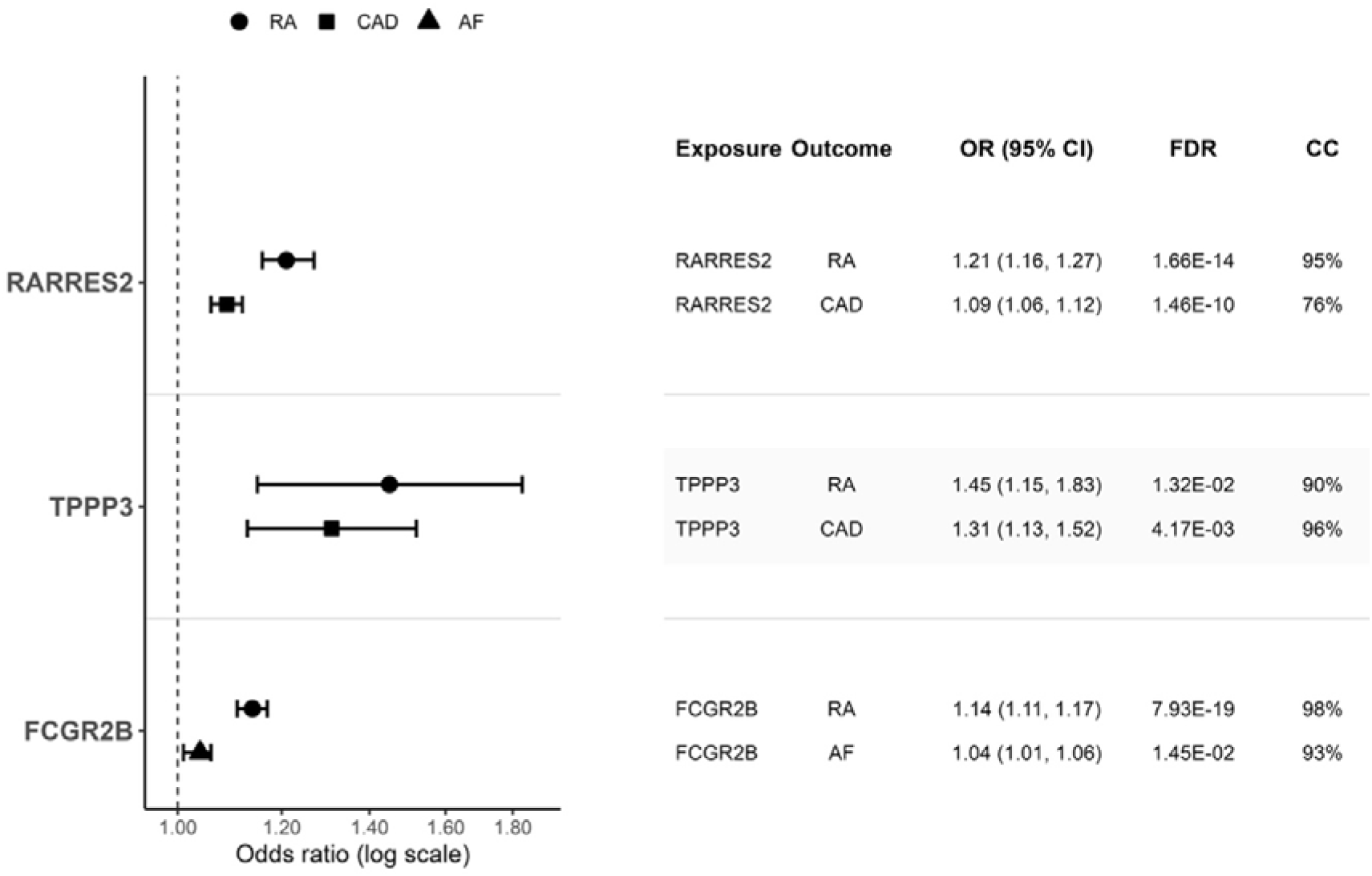
Forest plot illustrating the positive associations of genetically predicted RARRES2 and TPPP3 overlapping across RA and CAD, and FCGR2B in RA and AF.

**Figure 5.**
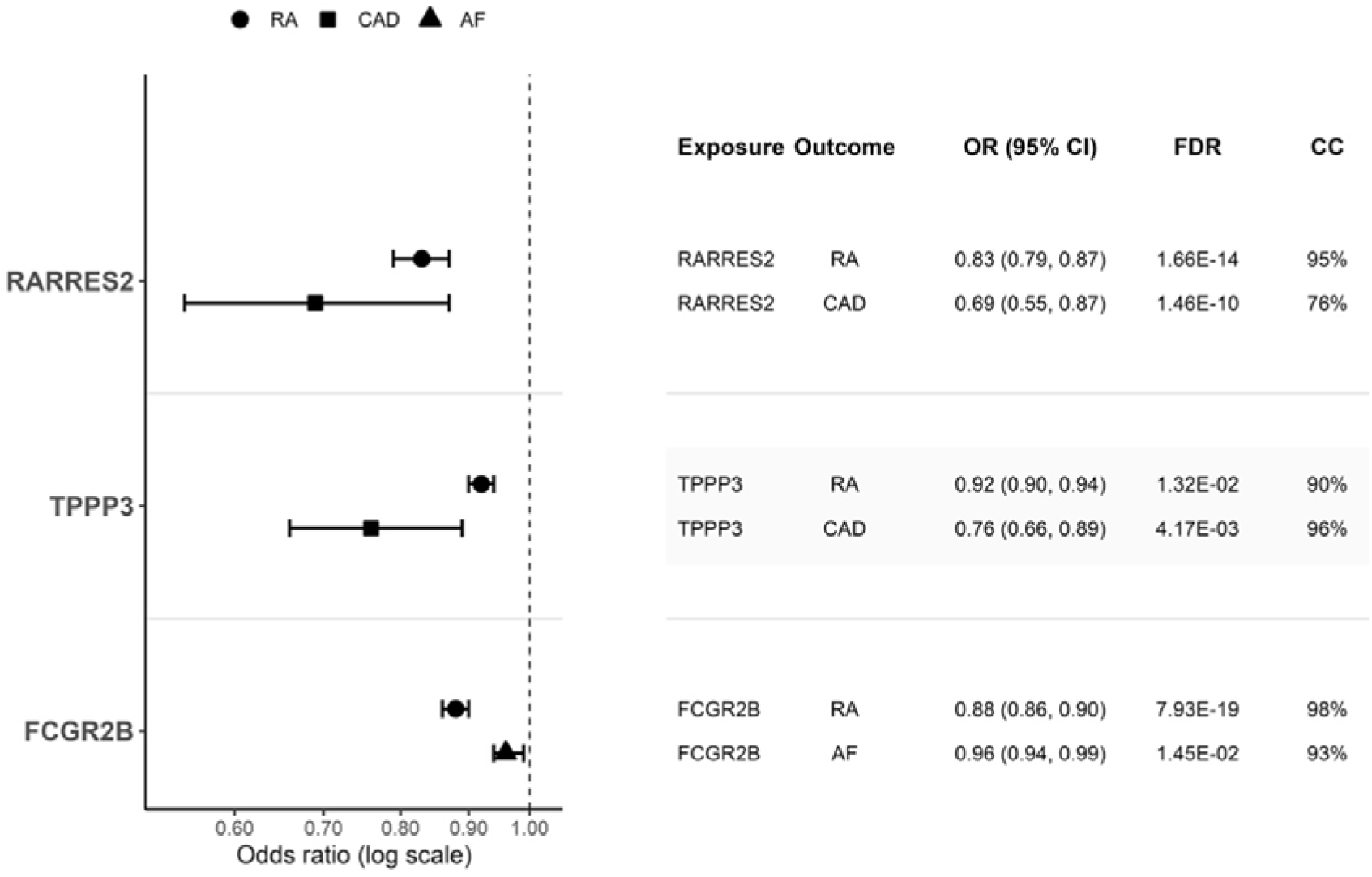
Forest plot illustrating the inverse associations of genetically predicted RARRES2 and TPPP3 overlapping across RA and CAD, and FCGR2B in RA and AF when being inhibited.

## Discussion

The findings from this study highlighted a group of RA-associated proteins with potential importance in both disease pathogenesis and cardiometabolic overlap. The results indicated that inhibition of RARRES2 and TPPP3 may reduce the risk of both and RA and CAD, whereas inhibition of FCGR2B may be associated with a lower risk of RA and AF. Furthermore, CD40, CD8A, TPPP3, CCL21, SNPX2, RARRES2, AKAP12, and GGT5 showed clear expression patterns within the local RA inflammatory environment. Druggability assessment showed that IFNGR2, IL6R, CD40, and FCGR2B were classified as Teir 1 whereas RARRES2, AKAP12, TPPP3, and SNX2 had limited available druggability data.

RARRES2, which encodes the secreted adipokine chemerin,^41^ is a biologically plausible shared risk factor for both RA and CAD. Chemerin is involved in immune regulation, inflammation and metabolic regulation,^41^ placing it at the intersection of systemic inflammation and cardiometabolic dysfunction. In RA, chemerin and its receptor (ChemR23) have been detected in synovial tissue, where the interaction of chemerin and its receptor can activate fibroblast-like synoviocytes and enhance inflammatory mediator expression.^42^ In cardiovascular disease, chemerin has been linked to endothelial dysfunction, monocyte adhesion, vascular inflammation, and atherosclerotic development.^43^ This might suggest that RARRES2 may represent a mechanistic bridge between chronic joint inflammation and accelerated atherosclerosis although whether it acts as a direct causal driver, a biomarker of inflammatory-metabolic burden or both remains unclear.

Some evidence suggests that TPPP3 may represent a biologically plausible link between RA and CAD. TPPP3 is an unstructured protein that promotes tubulin polymerisation and has been identified as a distinct marker of the developing musculoskeletal system. It is expressed in cells surrounding embryonic tendons, likely progenitors of the tendon sheath, epitenon and paratenon, and is also expressed in forming synovial joints, where its onset coincides with cavitation demonstrating a stage of joint differentiation. This developmental expression pattern supports the relevance of TPPP3 to joint-associated tissues^44^ such as RA which is a chronic autoimmune disease primarily affecting the synovium, where persistent inflammation leads to progressive cartilage and bone damage.^1^

Experimental evidence suggests that TPPP3 may promote oxidative endothelial injury through increased voltage-dependent anion channel 1 (VDAC1)-dependent pathway, as demonstrated in palmitic acid-treated human umbilical vein endothelial cells (HUVECs). Because endothelial dysfunction is a key early event in atherosclerosis, these findings provide biological plausibility for a potential role of TPPP3 in CAD-related vascular pathology. However, since the evidence derives from an in vitro endothelial cell model rather than coronary vascular tissue, atherosclerotic lesions, or in vivo disease systems, it should be interpreted as indirect support rather than proof of a CAD-specific mechanism. ^45, 46, 47^ Overall, TPPP3’s specific role in linking RA-related pathology to CAD requires further validation.

FCGR2B is a protein-coding gene that encodes Fc gamma receptor IIb, a low-affinity receptor for the Fc region of immunoglobulin G. It is mainly expressed on immune cells such as B cells, macrophages, and dendritic cells where it helps clear immune complexes and suppresses excessive immune activation. Because FCGR2B plays an important inhibitory role in immune regulation, this gene has been linked to autoimmune diseases such as systemic lupus erythematosus. Also, accumulating evidence supports its relevance in RA. Experimental studies in collagen-induced arthritis, a B cell-dependent model that shares features with RA have shown that FCGR2B deficiency increases collagen-specific immunoglobulin G levels and worsens arthritis severity whereas B cell-specific overexpression reduces both antibody titres and disease severity supporting a protective immunoregulatory role if this receptor in inflammatory joint disease.

In human RA, although FCGR2B 695T > C variant has not been consistently associated with RA susceptibility it has been linked to greater radiographic joint damage and altered dendritic cell responses to immune complexes indicating that FCGR2B may be more important in modulating disease progression and immune effector function than in determining RA onset itself. The present study suggests that FCGR2B is also linked to increased risk of RA. In relation to AF, existing evidence remains limited and not fully consistent; a proteome-wide Mendelian randomisation study reports that higher genetically predicted plasma levels of FCGR2B were associated with a lower risk of AF, in contrast to the findings from this study.^48, 49, 50, 51^ Further research is needed to clarify the shared biological mechanisms through which FCGR2B may contribute to both RA and AF.

One important strength of this study is the integration of large-scale blood-derived proteomic data with additional assessment in synovial tissue which is the main site of RA pathology. Although circulating proteins provide an accessible and informative source for biomarker discovery, blood-based signals may not always fully reflect disease-specific process occurring in the affected tissue.^12^ By examining whether the identified proteins were also supported in synovial tissue, this approach enhanced the disease relevance of the findings and increase confidence that the prioritised proteins were linked to RA-related biological mechanisms.

This study also provided additional insights into the management of RA. Several proteins linked to RA such as RARRES2, AKAP12, TPPP3, and SNX had limited available druggability data, which may reflect important gaps in current RA therapeutic knowledge. Addressing these gaps could contribute to improving treatment strategies and outcomes for patients with RA.

Several limitations should be considered when interpreting the findings. The analysis of RA-associated proteins in relation to cardiometabolic diseases was not performed in datasets specifically comprising individuals with both RA and the corresponding cardiometabolic condition. Therefore, the identified associations may not fully capture the biology of the comorbidity.

Although MR-based drug target analyses were performed, these estimates reflect lifelong genetically proxied effects and may differ from the effects observed following the clinical intervention which are influenced by treatment timing, dose, and duration.^18^.

Although the present results support RARRES2, TPPP3, and FCGR2B as potential inhibitory targets, the findings should be considered cautiously. The downstream biological effects of inhibiting these proteins are not yet fully understood, especially with respect to their influence on other diseases and physiological processes. It is possible that their inhibition could reduce the risk of RA and its related cardiovascular outcomes, but it may also introduce unintentional side effects in other contexts. For this reason, additional evaluation is required before translating these findings into therapeutic strategies. In particular, phenome-wide association analyses on comprehensive datasets could offer valuable insights into the wider phenotypic consequences of protein inhibition and help assess potential sides effects and trade-offs across different conditions.

## Conclusion

This study advances the understanding of RA by systematically linking RA-associated proteins to synovial relevance, therapeutic tractability, and cardiometabolic overlap. The findings show that while several proteins emerge as plausible therapeutic targets, the current knowledge remains incomplete with potentially important RA-related proteins lacking sufficient characterisation or druggability evidence. The targets identified across RA and the cardiometabolic diseases require more research to further understand their biology.

## Ethical approval

This study involved secondary analyses of publicly available GWAS summary statistics and UK Biobank summary statistics. The publicly available GWAS datasets were conducted in accordance with established ethical guidelines and approved by the relevant institutional review boards or ethics committees. UK Biobank has received ethical approval from the appropriate research ethics committee as well.

## Data availability

This research was conducted using data from the UK Biobank under approved application number 72723. UK Biobank data are available to researchers upon application and approval through the UK Biobank Access Management System. Further information on accessing UK Biobank data can be found at UK Biobank Access Management System https://ams.ukbiobank.ac.uk/ams/.

The following GWAS summary statistics analysed in this study are publicly available from their original sources:

1. RA data is a meta-analysis of Saevarsdottir S. et al. RA and Ishigaki et al. from the following links Saevarsdottir S. et al. (https://www.decode.com/summarydata/) Ishigaki RA (https://www.ebi.ac.uk/gwas/studies/GCST90132223),
2. Hypertension https://www.ebi.ac.uk/gwas/studies/GCST007610
3. CAD https://www.ebi.ac.uk/gwas/studies/GCST90132314
4. HF https://cvd.hugeamp.org/dinspector.html?dataset=GWAS_HERMES_eu&phenotype=HF
5. Stroke https://www.ebi.ac.uk/gwas/studies/GCST90104539
6. AF https://www.ebi.ac.uk/gwas/studies/GCST006414
7. T2DM https://t2d.hugeamp.org/dinspector.html?dataset=Mahajan2022_T2D_EU&phenotype=T2D
8. RA synovial tissue analysis was performed using the AMP RA Phase II single-cell platform available through Immunogenomics.io https://immunogenomics.io/ampra2/app/
9. Druggubility analysis was run using the work by Finan et al. https://www.science.org/doi/10.1126/scitranslmed.aag1166 as well as Open Targets Platform at https://platform.opentargets.org/downloads

